# A pragmatic model to forecast the COVID-19 epidemic in different countries and allowing for daily updates

**DOI:** 10.1101/2020.04.07.20056481

**Authors:** Carlos Nordt, Marcus Herdener

**Affiliations:** Psychiatric Hospital, University of Zurich, Zurich, Switzerland

## Abstract

Due to high infections rates and a high death toll of the COVID-19 pandemic, it is important to have daily updated forecasted estimates for the next weeks in order to allocate the scare resources as good as possible. We propose a pragmatic model to forecast the COVID-19 epidemic by applying a mixture normal distribution to open accessible WHO data. We specified a simple joint model on data from 20 countries with number of confirmed COVID-19 infections and number of COVID-19 deaths. We found that the duration of an epidemic wave (99% of total size) was usually between 45 – 48 days. Using data up to April 6, 2020, we found in six of 20 counties two waves, spaced between 21 and 47 days. In China and Korea the first wave was bigger, and in Denmark, Iran, Japan, and Sweden the second wave was stronger. Lag time between time trends in confirmed infections and time trends in deaths varied between 3.1 and 9.5 days. We obtained a good fit between observed and modelled data in almost all countries. In about halve of the countries the highest peak of the COVID-19 epidemic had been reached until April 6, 2020. Among the 20 countries, it is predicted that the USA will reach the highest numbers of confirmed infections (653 683 – 802 205) and number of deaths (36 591 – 53 286). Taken together, for many countries reasonable and up-to-date forecasting seems to be feasible. This method therefore bears a high potential for assisting decision makers to adjust the measures aiming at reducing the spread of the virus appropriately.

## Introduction

High infection rates and a high death toll due to the COVID-19 pandemic have been observed in many counties. Accordingly, various measures have been introduced to control the COVID- 19 epidemic, and these measures are accompanied by substantial restrictions both for individuals and the society as a whole in many countries. Therefore, it would be of great interest for good and updated estimates based on the most recent data on how the epidemic will develop in the near future, because this information is a prerequisite to adjust the measures limiting the spread of the virus appropriately.

It is well known that the COVID-19 originated in Hubei, a province of China, with first official recognition of a new virus in January 7, 2020. Since January 21, 2020 the World Health Organization (WHO) publishes a daily Situation Reports of the Coronavirus disease 2019 (https://www.who.int/emergencies/diseases/novel-coronavirus-2019/situation-reports). This daily report includes the number of confirmed infections and number of deaths by country. Although the incidence rates for infections and the ratio between number of infections and number of deaths varies substantially between countries, there are strong similarities between the time trends on country level. Evidence so far, based on the existing data sources, suggests that the (first) wave of the epidemic can be approximated by a unimodal distribution.

Therefore, we propose a pragmatic model that allows for daily updated forecasts of the regional spread of the pandemic, and that is based on the following assumptions: the development of number of daily confirmed infections as well as the number of daily deaths can be described by a normal distribution, where there is a certain time lag of the latter. Obviously, the start of the epidemic as well as the size of the epidemic are country specific. But as there is a similar pattern how governments react to the COVID-19 epidemic (first trying to ignoring them, then introducing similar steps of measures to control the epidemic), we assume that the duration of the epidemic waves are also similar across countries. In those countries, where the data indicate two waves (i.e. China, Denmark, Iran, Japan, Korea, and Sweden) we applied a mixture of two normal distributions (where the proportional size of the waves and the time lag between the maximum peak are estimated, but standard deviation is assumed to be identical). Additionally, we use only data from the WHO reports if the cumulative number of confirmed COVID-19 infections were 100 cases or higher, as this probably indicates onset of local transmission.

As for some countries WHO data are not updated every day and given that it is almost impossible to collect such data with delays of less than 24 hours, we used moving averages with a time window of 5 days for analysis in order to address such ‘data spikes’ that are most probably artificial and due to the collection and reporting process. There was also a change in definition how to report confirmed cases in China resulting in an extreme peak in the WHO data. Moreover, it is also probable that availability of tests and installing routes of information procedures may change over time in a country. Despite these biases, we think that our simple and pragmatic model will result in useful forecasts that can be updated every day due to its simplicity, and that can easily be applied on data on a national or regional level.

For analysis, we used data of 20 countries and applied a joint model, with number of confirmed infections and number of deaths as dependent variable. We used a binomial distribution and accounted for model deviations by variance-covariance matrix adjustment. On April 6, 2020, the model could use 1 414 data points to estimate 76 parameters in total.

## Results

The application of our pragmatic COVID-19 forecast model using data of twenty countries lead to the following results (see also Figure 1 and Figure 2):

**Figure 1:**
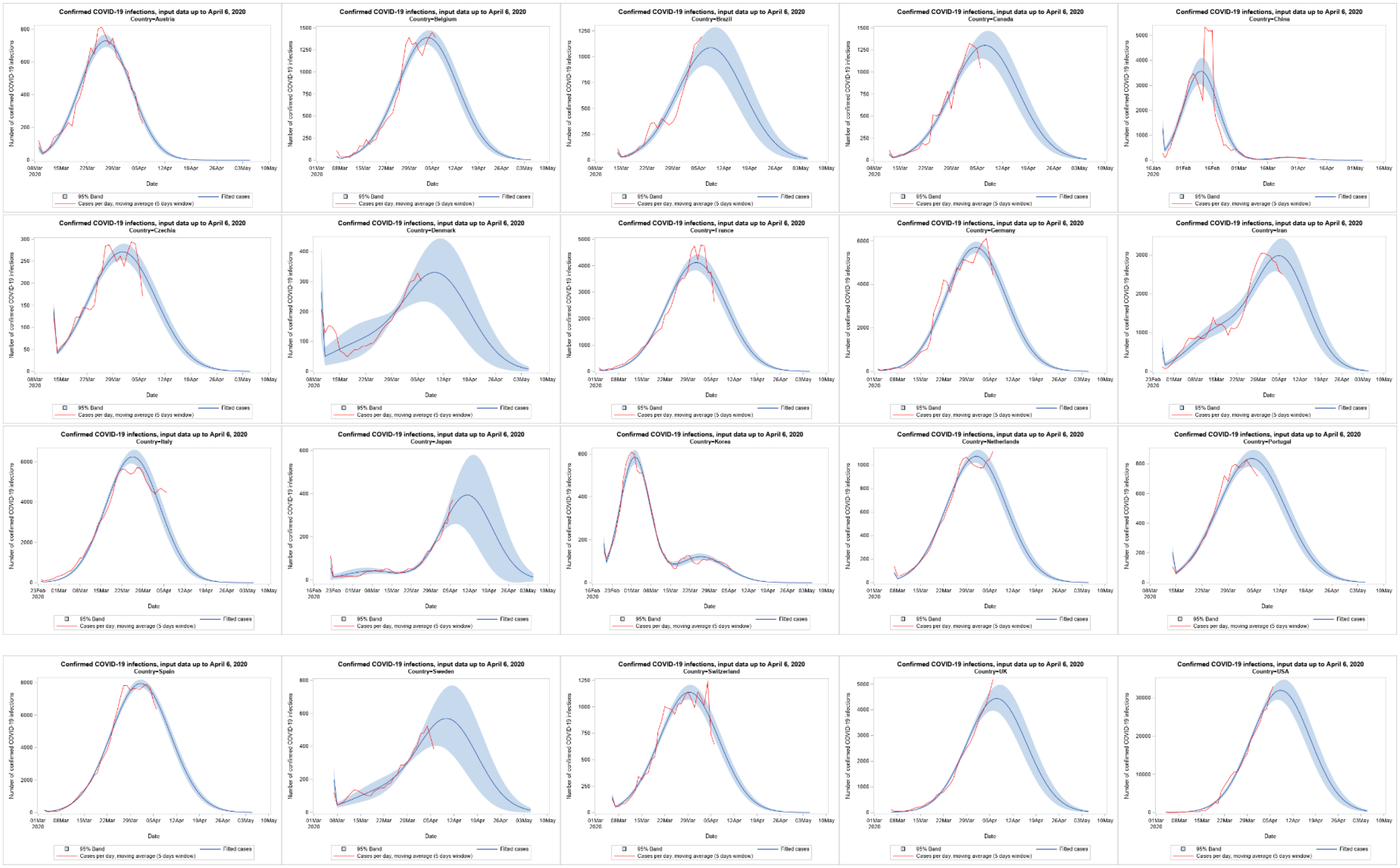
Observed (red line) and predicted number of confirmed COVID-19 infection in 20 countries, using data from April 6, 2020

**Figure 2:**
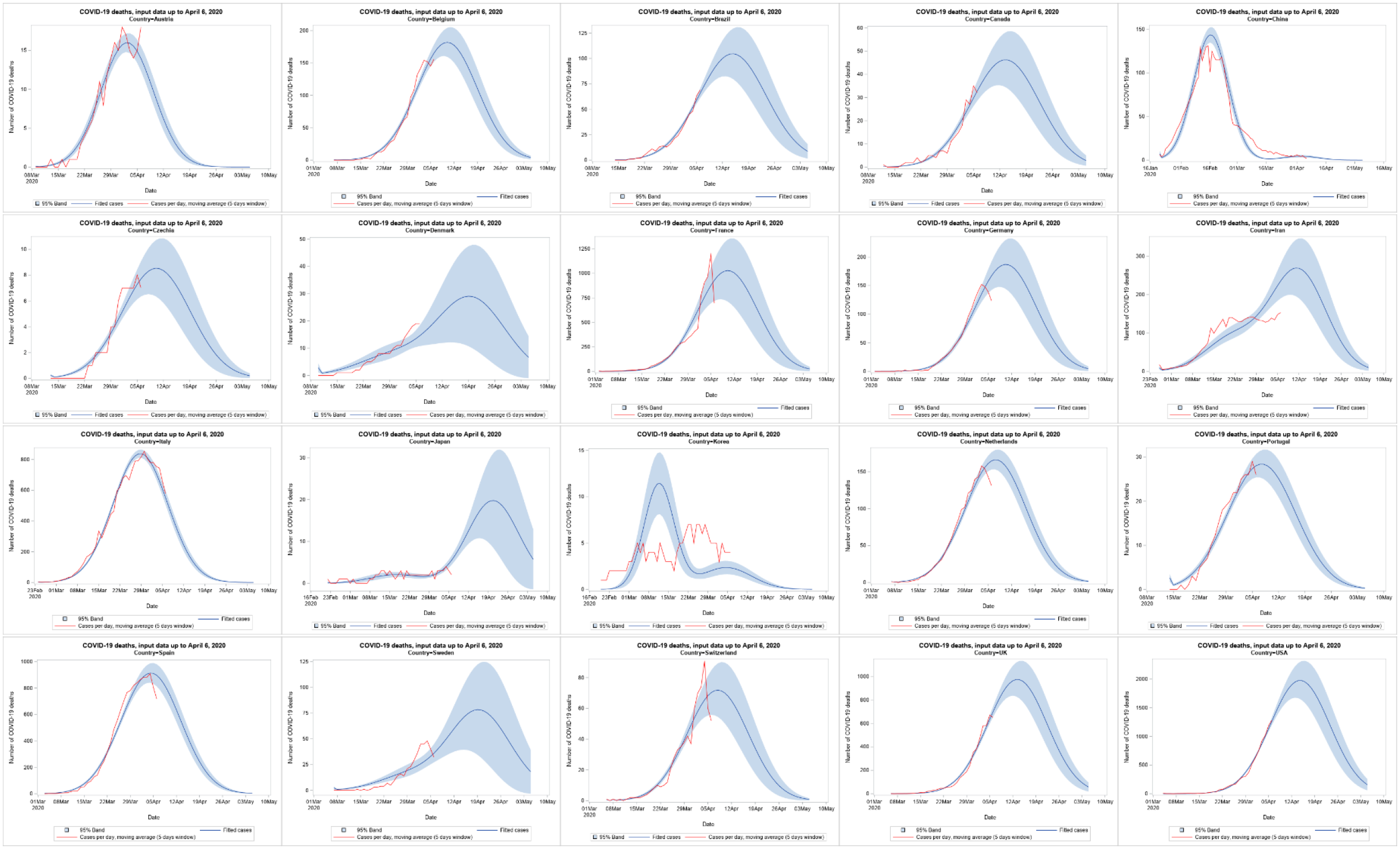
Observed (red line) and predicted number of COVID-19 deaths in 20 countries, using data from April 6, 2020

The standard distribution of COVID-19 epidemics was estimated to be between 8.8 and 9.4 days, meaning that 99% of confirmed COVID-19 cases or deaths within a given region or country will occur between 45 and 48 days (Table 1, third last line). There are two exceptions to the standard distribution: the first COVID-19 wave in Korea was estimated to last 26.6 – 28.7 days and in Austria the duration of the COVID-19 wave was estimated to be between 34.4 and 38.1 days.

**Table 1:** Derived estimates for the 20 countries.

| **Label** | **Estimate** | **Standard Error** | **DF** | **t Value** | **Pr > \|t\|** | **Alpha** | **Lower** | **Upper** |
| --- | --- | --- | --- | --- | --- | --- | --- | --- |
| **Peak of 1st. wave China (day of 2020)** | 40.5137 | 0.4974 | 76 | 81.46 | <.0001 | 0.05 | 39.5232 | 41.5043 |
| **Peak of 2nd. wave China (day of 2020)** | 87.1299 | 0.4732 | 76 | 184.13 | <.0001 | 0.05 | 86.1874 | 88.0724 |
| **Time between peaks China (days)** | 46.6162 | 0.5762 | 76 | 80.91 | <.0001 | 0.05 | 45.4686 | 47.7637 |
| **Size of 1st. wave China (%)** | 96.9532 | 0.2337 | 76 | 414.94 | <.0001 | 0.05 | 96.4878 | 97.4186 |
| **Total confirmed China** | 83700 | 5473.61 | 76 | 15.29 | <.0001 | 0.05 | 72798 | 94602 |
| **Total deaths China** | 3366.92 | 100.25 | 76 | 33.59 | <.0001 | 0.05 | 3167.25 | 3566.59 |
| **Ratio deaths per confirmed China (%)** | 4.0226 | 0.3099 | 76 | 12.98 | <.0001 | 0.05 | 3.4054 | 4.6399 |
| **Time lag of deaths vr. confirmed China (days)** | 6.3101 | 0.3906 | 76 | 16.15 | <.0001 | 0.05 | 5.5322 | 7.0881 |
| **Peak of 1st. wave Korea (day of 2020)** | 61.6431 | 0.09430 | 76 | 653.67 | <.0001 | 0.05 | 61.4552 | 61.8309 |
| **Peak of 2nd. wave Korea (day of 2020)** | 85.4521 | 0.4740 | 76 | 180.28 | <.0001 | 0.05 | 84.5081 | 86.3962 |
| **Time between peaks Korea (days)** | 23.8091 | 0.4495 | 76 | 52.96 | <.0001 | 0.05 | 22.9137 | 24.7044 |
| **Size of 1st. wave Korea (%)** | 73.9857 | 0.8099 | 76 | 91.36 | <.0001 | 0.05 | 72.3727 | 75.5986 |
| **Total confirmed Korea** | 10564 | 159.01 | 76 | 66.44 | <.0001 | 0.05 | 10248 | 10881 |
| **Total deaths Korea** | 206.46 | 29.7374 | 76 | 6.94 | <.0001 | 0.05 | 147.24 | 265.69 |
| **Ratio deaths per confirmed Korea (%)** | 1.9544 | 0.2856 | 76 | 6.84 | <.0001 | 0.05 | 1.3855 | 2.5233 |
| **Time lag of deaths vr. confirmed Korea (days)** | 9.5248 | 0.6973 | 76 | 13.66 | <.0001 | 0.05 | 8.1360 | 10.9136 |
| **Peak of 1st. wave Japan (day of 2020)** | 67.1781 | 1.9100 | 76 | 35.17 | <.0001 | 0.05 | 63.3740 | 70.9821 |
| **Peak of 2nd. wave Japan (day of 2020)** | 101.66 | 1.6834 | 76 | 60.39 | <.0001 | 0.05 | 98.3085 | 105.01 |
| **Time between peaks Japan (days)** | 34.4833 | 1.1954 | 76 | 28.85 | <.0001 | 0.05 | 32.1024 | 36.8641 |
| **Size of 1st. wave Japan (%)** | 9.4846 | 1.5759 | 76 | 6.02 | <.0001 | 0.05 | 6.3460 | 12.6232 |
| **Total confirmed Japan** | 9890.41 | 1883.39 | 76 | 5.25 | <.0001 | 0.05 | 6139.33 | 13641 |
| **Total deaths Japan** | 496.61 | 129.17 | 76 | 3.84 | 0.0002 | 0.05 | 239.34 | 753.88 |
| **Ratio deaths per confirmed Japan (%)** | 5.0211 | 0.6177 | 76 | 8.13 | <.0001 | 0.05 | 3.7909 | 6.2513 |

**Table 1: Derived estimates for the 20 countries**
| **Label** | **Estimate** | **Standard Error** | **DF** | **t Value** | **Pr > \|t\|** | **Alpha** | **Lower** | **Upper** |
| --- | --- | --- | --- | --- | --- | --- | --- | --- |
| **Time lag of deaths vr. confirmed Japan (days)** | 9.5248 | 0.6973 | 76 | 13.66 | <.0001 | 0.05 | 8.1360 | 10.9136 |
| **Peak of 1st. wave Iran (day of 2020)** | 74.4012 | 1.1258 | 76 | 66.09 | <.0001 | 0.05 | 72.1590 | 76.6434 |
| **Peak of 2nd. wave Iran (day of 2020)** | 95.9140 | 0.8768 | 76 | 109.39 | <.0001 | 0.05 | 94.1677 | 97.6603 |
| **Time between peaks Iran (days)** | 21.5128 | 1.0362 | 76 | 20.76 | <.0001 | 0.05 | 19.4491 | 23.5765 |
| **Size of 1st. wave Iran (%)** | 24.8075 | 1.5933 | 76 | 15.57 | <.0001 | 0.05 | 21.6342 | 27.9807 |
| **Total confirmed Iran** | 88479 | 4152.15 | 76 | 21.31 | <.0001 | 0.05 | 80209 | 96749 |
| **Total deaths Iran** | 7974.96 | 928.15 | 76 | 8.59 | <.0001 | 0.05 | 6126.39 | 9823.53 |
| **Ratio deaths per confirmed Iran (%)** | 9.0134 | 0.7543 | 76 | 11.95 | <.0001 | 0.05 | 7.5110 | 10.5158 |
| **Time lag of deaths vr. confirmed Iran (days)** | 6.3101 | 0.3906 | 76 | 16.15 | <.0001 | 0.05 | 5.5322 | 7.0881 |
| **Peak of wave Italy (day of 2020)** | 85.0982 | 0.3970 | 76 | 214.34 | <.0001 | 0.05 | 84.3075 | 85.8890 |
| **Total confirmed Italy** | 142275 | 4579.09 | 76 | 31.07 | <.0001 | 0.05 | 133155 | 151395 |
| **Total deaths Italy** | 19054 | 394.51 | 76 | 48.30 | <.0001 | 0.05 | 18268 | 19840 |
| **Ratio deaths per confirmed Italy (%)** | 13.3923 | 0.2607 | 76 | 51.37 | <.0001 | 0.05 | 12.8731 | 13.9115 |
| **Time lag of deaths vr. confirmed Italy (days)** | 3.1250 | 0.2408 | 76 | 12.98 | <.0001 | 0.05 | 2.6453 | 3.6046 |
| **Peak of 1st. wave Denmark (day of 2020)** | 79.7005 | 1.3762 | 76 | 57.91 | <.0001 | 0.05 | 76.9595 | 82.4415 |
| **Peak of 2nd. wave Denmark (day of 2020)** | 100.50 | 1.2904 | 76 | 77.89 | <.0001 | 0.05 | 97.9341 | 103.07 |
| **Time between peaks Denmark (days)** | 20.8037 | 1.2571 | 76 | 16.55 | <.0001 | 0.05 | 18.3000 | 23.3075 |
| **Size of 1st. wave Denmark (%)** | 20.0012 | 4.2176 | 76 | 4.74 | <.0001 | 0.05 | 11.6010 | 28.4013 |
| **Total confirmed Denmark** | 9218.81 | 992.00 | 76 | 9.29 | <.0001 | 0.05 | 7243.07 | 11195 |
| **Total deaths Denmark** | 810.67 | 199.40 | 76 | 4.07 | 0.0001 | 0.05 | 413.53 | 1207.81 |
| **Ratio deaths per confirmed Denmark (%)** | 8.7937 | 1.5637 | 76 | 5.62 | <.0001 | 0.05 | 5.6793 | 11.9080 |
| **Time lag of deaths vr. confirmed Denmark (days)** | 9.5248 | 0.6973 | 76 | 13.66 | <.0001 | 0.05 | 8.1360 | 10.9136 |
| **Peak of wave Switzerland (day of 2020)** | 88.8916 | 0.3520 | 76 | 252.50 | <.0001 | 0.05 | 88.1905 | 89.5928 |

Table 1: Derived estimates for the 20 countries
| Label | Estimate | Standard Error | DF | t Value | Pr > \|t\| | Alpha | Lower | Upper |
| --- | --- | --- | --- | --- | --- | --- | --- | --- |
| **Total confirmed Switzerland** | 25869 | 817.89 | 76 | 31.63 | <.0001 | 0.05 | 24240 | 27498 |
| **Total deaths Switzerland** | 1634.47 | 205.52 | 76 | 7.95 | <.0001 | 0.05 | 1225.14 | 2043.80 |
| **Ratio deaths per confirmed Switzerland (%)** | 6.3183 | 0.6732 | 76 | 9.39 | <.0001 | 0.05 | 4.9775 | 7.6592 |
| **Time lag of deaths vr. confirmed Switzerland (days)** | 9.5248 | 0.6973 | 76 | 13.66 | <.0001 | 0.05 | 8.1360 | 10.9136 |
| **Peak of wave Austria (day of 2020)** | 86.5209 | 0.1816 | 76 | 476.51 | <.0001 | 0.05 | 86.1592 | 86.8825 |
| **Total confirmed Austria** | 12860 | 256.90 | 76 | 50.06 | <.0001 | 0.05 | 12349 | 13372 |
| **Total deaths Austria** | 281.84 | 10.2544 | 76 | 27.48 | <.0001 | 0.05 | 261.41 | 302.26 |
| **Ratio deaths per confirmed Austria (%)** | 2.1916 | 0.08491 | 76 | 25.81 | <.0001 | 0.05 | 2.0225 | 2.3607 |
| **Time lag of deaths vr. confirmed Austria (days)** | 6.3101 | 0.3906 | 76 | 16.15 | <.0001 | 0.05 | 5.5322 | 7.0881 |
| **Peak of 1st. wave Sweden (day of 2020)** | 79.7005 | 1.3762 | 76 | 57.91 | <.0001 | 0.05 | 76.9595 | 82.4415 |
| **Peak of 2nd. wave Sweden (day of 2020)** | 100.50 | 1.2904 | 76 | 77.89 | <.0001 | 0.05 | 97.9341 | 103.07 |
| **Time between peaks Sweden (days)** | 20.8037 | 1.2571 | 76 | 16.55 | <.0001 | 0.05 | 18.3000 | 23.3075 |
| **Size of 1st. wave Sweden (%)** | 16.0229 | 2.5612 | 76 | 6.26 | <.0001 | 0.05 | 10.9219 | 21.1239 |
| **Total confirmed Sweden** | 15201 | 2052.30 | 76 | 7.41 | <.0001 | 0.05 | 11113 | 19288 |
| **Total deaths Sweden** | 2092.39 | 529.98 | 76 | 3.95 | 0.0002 | 0.05 | 1036.85 | 3147.92 |
| **Ratio deaths per confirmed Sweden (%)** | 13.7653 | 2.1831 | 76 | 6.31 | <.0001 | 0.05 | 9.4171 | 18.1134 |
| **Time lag of deaths vr. confirmed Sweden (days)** | 9.5248 | 0.6973 | 76 | 13.66 | <.0001 | 0.05 | 8.1360 | 10.9136 |
| **Peak of wave Germany (day of 2020)** | 91.2758 | 0.3769 | 76 | 242.16 | <.0001 | 0.05 | 90.5251 | 92.0265 |
| **Total confirmed Germany** | 129593 | 3620.72 | 76 | 35.79 | <.0001 | 0.05 | 122382 | 136805 |
| **Total deaths Germany** | 4248.38 | 511.64 | 76 | 8.30 | <.0001 | 0.05 | 3229.36 | 5267.41 |
| **Ratio deaths per confirmed Germany (%)** | 3.2782 | 0.3558 | 76 | 9.21 | <.0001 | 0.05 | 2.5697 | 3.9868 |
| **Peak of wave France (day of 2020)** | 91.0830 | 0.3683 | 76 | 247.28 | <.0001 | 0.05 | 90.3494 | 91.8166 |
| **Total confirmed France** | 93709 | 3519.89 | 76 | 26.62 | <.0001 | 0.05 | 86698 | 100719 |
| **Total deaths France** | 23379 | 3724.26 | 76 | 6.28 | <.0001 | 0.05 | 15962 | 30797 |

Table 1: Derived estimates for the 20 countries
| Label | Estimate | Standard Error | DF | t Value | Pr > \|t\| | Alpha | Lower | Upper |
| --- | --- | --- | --- | --- | --- | --- | --- | --- |
| Ratio deaths per confirmed France (%) | 24.9489 | 3.4918 | 76 | 7.15 | <.0001 | 0.05 | 17.9945 | 31.9034 |
| Peak of wave Spain (day of 2020) | 91.6253 | 0.2396 | 76 | 382.37 | <.0001 | 0.05 | 91.1481 | 92.1026 |
| Total confirmed Spain | 180790 | 3544.84 | 76 | 51.00 | <.0001 | 0.05 | 173730 | 187850 |
| Total deaths Spain | 20770 | 868.45 | 76 | 23.92 | <.0001 | 0.05 | 19041 | 22500 |
| Ratio deaths per confirmed Spain (%) | 11.4887 | 0.3682 | 76 | 31.20 | <.0001 | 0.05 | 10.7554 | 12.2220 |
| Peak of wave Netherlands (day of 2020) | 91.4433 | 0.3503 | 76 | 261.04 | <.0001 | 0.05 | 90.7456 | 92.1410 |
| Total confirmed Netherlands | 24449 | 801.38 | 76 | 30.51 | <.0001 | 0.05 | 22853 | 26045 |
| Total deaths Netherlands | 3781.55 | 182.75 | 76 | 20.69 | <.0001 | 0.05 | 3417.58 | 4145.52 |
| Ratio deaths per confirmed Netherlands (%) | 15.4669 | 0.8794 | 76 | 17.59 | <.0001 | 0.05 | 13.7154 | 17.2183 |
| Peak of wave Czechia (day of 2020) | 91.0802 | 0.3636 | 76 | 250.52 | <.0001 | 0.05 | 90.3561 | 91.8043 |
| Total confirmed Czechia | 6172.80 | 221.94 | 76 | 27.81 | <.0001 | 0.05 | 5730.77 | 6614.82 |
| Total deaths Czechia | 193.92 | 26.2781 | 76 | 7.38 | <.0001 | 0.05 | 141.58 | 246.26 |
| Ratio deaths per confirmed Czechia (%) | 3.1415 | 0.3674 | 76 | 8.55 | <.0001 | 0.05 | 2.4098 | 3.8733 |
| Peak of wave Belgium (day of 2020) | 94.1727 | 0.4382 | 76 | 214.92 | <.0001 | 0.05 | 93.3000 | 95.0454 |
| Total confirmed Belgium | 31644 | 1166.44 | 76 | 27.13 | <.0001 | 0.05 | 29321 | 33967 |
| Total deaths Belgium | 4133.11 | 291.94 | 76 | 14.16 | <.0001 | 0.05 | 3551.66 | 4714.56 |
| Ratio deaths per confirmed Belgium (%) | 13.0612 | 0.9160 | 76 | 14.26 | <.0001 | 0.05 | 11.2369 | 14.8855 |
| Peak of wave Portugal (day of 2020) | 94.8623 | 0.4362 | 76 | 217.45 | <.0001 | 0.05 | 93.9935 | 95.7312 |
| Total confirmed Portugal | 19012 | 660.44 | 76 | 28.79 | <.0001 | 0.05 | 17697 | 20327 |
| Total deaths Portugal | 646.27 | 37.0682 | 76 | 17.43 | <.0001 | 0.05 | 572.44 | 720.09 |
| Ratio deaths per confirmed Portugal (%) | 3.3992 | 0.1005 | 76 | 33.81 | <.0001 | 0.05 | 3.1990 | 3.5995 |
| Peak of wave UK (day of 2020) | 97.6017 | 0.6054 | 76 | 161.21 | <.0001 | 0.05 | 96.3958 | 98.8075 |
| Total confirmed UK | 100992 | 6814.55 | 76 | 14.82 | <.0001 | 0.05 | 87420 | 114565 |
| Total deaths UK | 22149 | 2006.92 | 76 | 11.04 | <.0001 | 0.05 | 18152 | 26146 |
| Ratio deaths per confirmed UK (%) | 21.9312 | 1.5390 | 76 | 14.25 | <.0001 | 0.05 | 18.8660 | 24.9965 |

**Table 1: Derived estimates for the 20 countries**
| **Label** | **Estimate** | **Standard Error** | **DF** | **t Value** | **Pr > \|t\|** | **Alpha** | **Lower** | **Upper** |
| --- | --- | --- | --- | --- | --- | --- | --- | --- |
| **Peak of wave Brazil (day of 2020)** | 98.9297 | 0.8591 | 76 | 115.15 | <.0001 | 0.05 | 97.2186 | 100.64 |
| **Total confirmed Brazil** | 24739 | 2367.54 | 76 | 10.45 | <.0001 | 0.05 | 20023 | 29454 |
| **Total deaths Brazil** | 2378.02 | 307.72 | 76 | 7.73 | <.0001 | 0.05 | 1765.14 | 2990.89 |
| **Ratio deaths per confirmed Brazil (%)** | 9.6126 | 0.7593 | 76 | 12.66 | <.0001 | 0.05 | 8.1002 | 11.1250 |
| **Peak of wave USA (day of 2020)** | 98.7798 | 0.5368 | 76 | 184.02 | <.0001 | 0.05 | 97.7107 | 99.8490 |
| **Total confirmed USA** | 727944 | 37286 | 76 | 19.52 | <.0001 | 0.05 | 653683 | 802205 |
| **Total deaths USA** | 44939 | 4191.08 | 76 | 10.72 | <.0001 | 0.05 | 36591 | 53286 |
| **Ratio deaths per confirmed USA (%)** | 6.1734 | 0.4051 | 76 | 15.24 | <.0001 | 0.05 | 5.3665 | 6.9802 |
| **Peak of wave Canada (day of 2020)** | 97.7030 | 0.6031 | 76 | 161.99 | <.0001 | 0.05 | 96.5017 | 98.9042 |
| **Total confirmed Canada** | 29651 | 1905.84 | 76 | 15.56 | <.0001 | 0.05 | 25855 | 33447 |
| **Total deaths Canada** | 1052.24 | 140.90 | 76 | 7.47 | <.0001 | 0.05 | 771.61 | 1332.87 |
| **Ratio deaths per confirmed Canada (%)** | 3.5487 | 0.2969 | 76 | 11.95 | <.0001 | 0.05 | 2.9573 | 4.1401 |
| **Duration of COVID-19 wave (99%) in days** | 46.7877 | 0.7668 | 76 | 61.02 | <.0001 | 0.05 | 45.2605 | 48.3150 |
| **Duration of first COVID-19 wave (99%) in Korea in days** | 27.6039 | 0.5267 | 76 | 52.41 | <.0001 | 0.05 | 26.5550 | 28.6529 |
| **Duration of COVID-19 wave (99%) in Austria in days** | 36.2422 | 0.9112 | 76 | 39.78 | <.0001 | 0.05 | 34.4275 | 38.0569 |

The time trends in number of deaths follows the trends in the number of confirmed infections and can be described by lag time, which differs between countries. This lag time was lowest (3.1 days) in Italy, Portugal, and Spain; medium (6.3 days) in Austria, Belgium, Brazil, Canada, China, Iran, Netherlands, UK, and USA; highest (9.5 days) in Czechia, Denmark, France, Germany, Japan, Korea, Sweden, and Switzerland.

To enhance comparisons of the timing of the estimated peaks in number of confirmed infections, we present these as number of day in 2020 (range 1 to 366; for example day 32 in 2020 is Feb 1). In China the first peak was estimated at day 40 (accounting for about 97% of total COVID-19 epidemic in China), and a small second peak at day 87 (accounting for 3% of total). The estimated total of the whole epidemic in China of 83 700 confirmed infections is close to the 83 005 confirmed infections reported by the WHO on April 6, 2020 for China. Also the estimated total of the whole epidemic in China of 3 367 deaths is close to the 3 340 reported deaths by the WHO. That is due to the fact that the model indicates that the COVID-19 epidemic in China is close to its end (at least regarding the current wave), and thus differences between model estimates and observed cases could only stem from misspecification of the model. The lower confidence interval of the model is with 72 789 confirmed infections about 10 000 cases lower than the observed numbers. This difference is due to the fact that the change in definition of how confirmed infections are counted is not accounted for in our model.

Besides China, there are five other countries with two peaks: Korea with a first peak at day 62 (74% of total) and a second peak at day 85 (26% of total); Japan with a first peak on day 67 (9% of total) and a second peak on day 102 (91% of total); Iran with a first peak on day 74 (25% of total) and a second peak on day 96 (75%); the peaks of the waves in Denmark and Sweden where assumed to be identical, with first peak on day 80 (20% of total in Denmark and 16% of total in Sweden), and a second peak on day 101.

For all other 14 countries only one peak could be observed according to the currently available data. Italy had its peak on day 85, Austria had its peak on day 87, Switzerland on day 89, Czechia, France, Germany, and Netherlands all on day 91, Spain on day 92, Belgium on day 94, Portugal on day 95, UK and Canada on day 98, Brazil and USA on day 99.

Among these 20 countries, it is predicted that the USA will reach the highest numbers (653 683 – 802 205) of confirmed infections and number of deaths (36 591 – 53 286).

Despite the fact that confidence intervals of our model are rather wide, we believe that our forecast can be helpful to inform decision makers. For example, for Switzerland the known cases on April 6, 2020 are 21 065 confirmed infections, and the model predicts a total between 24 240 and 27 498 confirmed infections; the observed number of deaths was 715, and the model forecasted between 1 225 and 2 044 deaths. Therefore, we predict that the majority of people who will get infected during this (first) epidemic wave are already infected.

If the model is estimated with the data provided by the Swiss cantons (www.corona-data.ch), using all cantonal data and accounting for days without new published data and setting the standard deviation of the normal distribution to 9, the estimated total number of confirmed COVID-19 infections is with 22 680 – 27 167 almost identical to the model using WHO data (see Figure 3 and Table 2 for Swiss cantons).

**Table 2:** Observed and predicted total number of confirmed COVID-19 infection in the 26 cantons of Switzerland, using data from April 6, 2020

|  | AG | AI | AR | BE | BL | BS | FR | GE | GL | GR | JU | LU | NE | NW | OW | SG | SH | SO | SZ | TG | TI | UR | VD | VS | ZG | ZH | CH |
| --- | --- | --- | --- | --- | --- | --- | --- | --- | --- | --- | --- | --- | --- | --- | --- | --- | --- | --- | --- | --- | --- | --- | --- | --- | --- | --- | --- |
| Observed until April 6, 2020 | 727 | 21 | 69 | 1173 | 682 | 803 | 669 | 3439 | 63 | 646 | 160 | 497 | 459 | 86 | 60 | 532 | 49 | 261 | 178 | 219 | 2546 | 67 | 4115 | 1400 | 152 | 2590 | 21663 |
| Predicted total | 819 | 24 | 77 | 1329 | 770 | 911 | 786 | 4023 | 70 | 763 | 182 | 563 | 537 | 95 | 66 | 600 | 57 | 295 | 203 | 250 | 2895 | 76 | 4811 | 1600 | 173 | 2948 | 24923 |
| 95% lower | 662 | 10 | 54 | 1042 | 536 | 773 | 646 | 3342 | 47 | 572 | 128 | 455 | 444 | 70 | 44 | 337 | 32 | 231 | 143 | 191 | 2230 | 48 | 3826 | 1389 | 130 | 2388 | 22680 |
| 95% upper | 977 | 37 | 100 | 1617 | 1005 | 1049 | 925 | 4703 | 93 | 954 | 236 | 671 | 631 | 120 | 88 | 862 | 82 | 358 | 264 | 309 | 3560 | 104 | 5796 | 1812 | 216 | 3507 | 27166 |

**Figure 3:**
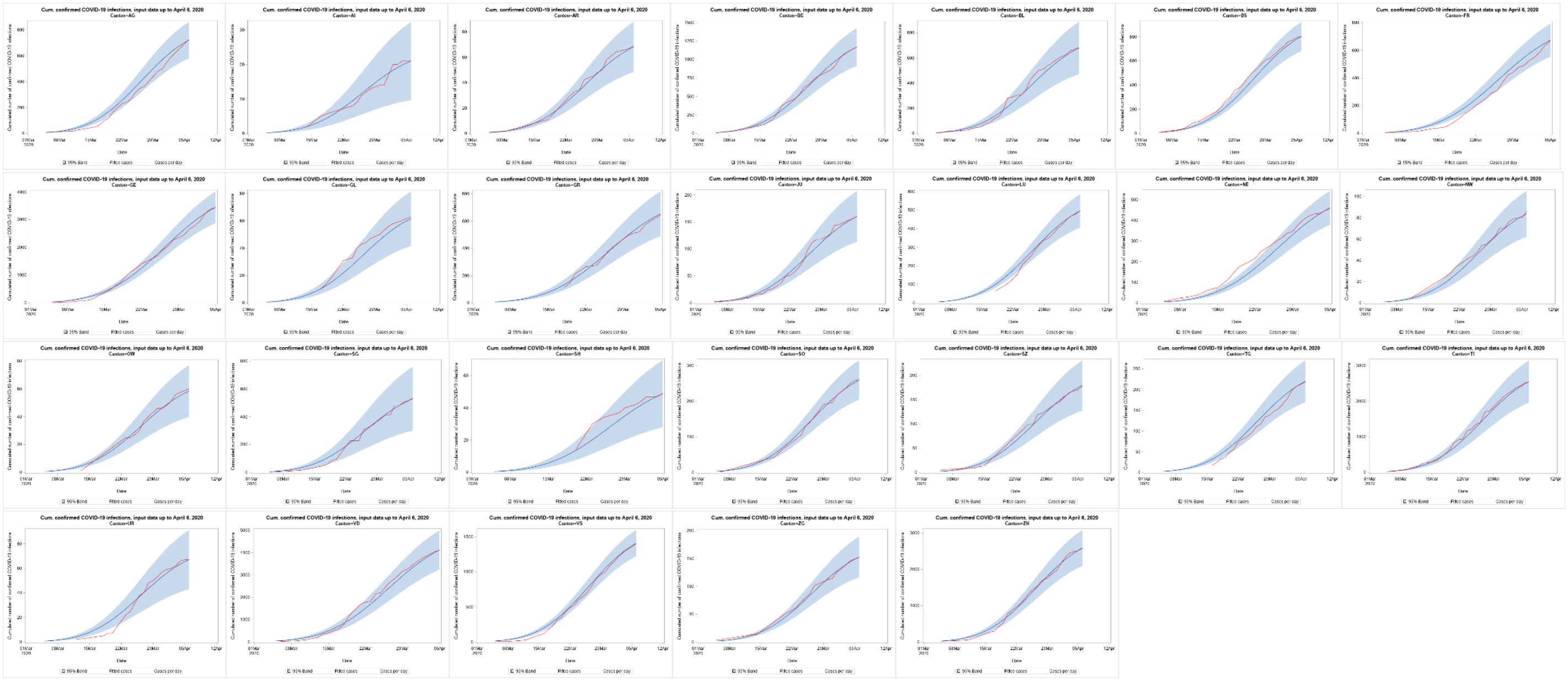
Observed and model fitted cumulative number of confirmed COVID-19 infection in the 26 cantons of Switzerland, using data from April 6, 2020

## Methods

The model was estimated wit SAS 9.4M6 using PROC NLMIXED. The input variables are “date” for the date, an indicator variable named “dead” that is zero if “cases” indicate new confirmed infections, and one if “cases” indicate new number of deaths. We show the syntax here for China solely, as otherwise the syntax is quite extensive.

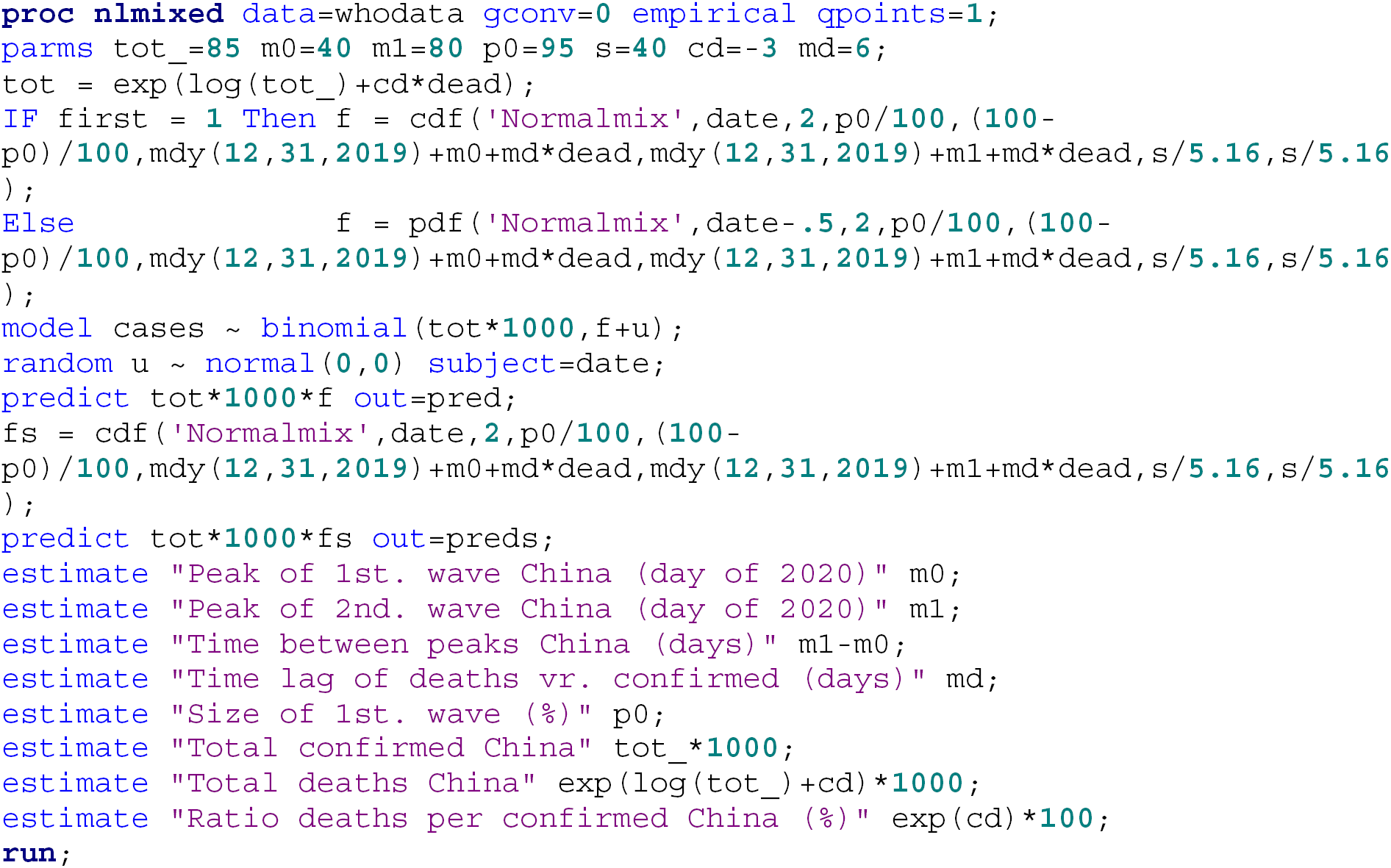

## Data Availability

We used only data from data bases that are open access: https://www.who.int/emergencies/diseases/novel-coronavirus-2019/situation-reports
and 
www.corona-data.ch

